# Expanding *Kelch13* artemisinin resistance markers are associated with higher malaria incidence in Uganda

**DOI:** 10.64898/2026.10.19.26365887

**Authors:** Stephanie van Wyk, Jonathan Gwasupika, Lovemore Mapahla, Victor Asua, Lucinda E Harrison, Moses R. Kamya, Joaniter Nankabirwa, Karen I Barnes

## Abstract

Artemisinin partial resistance (ART-R)-associated *Plasmodium falciparum Kelch13* mutations are expanding across Africa. However, their population-level consequences remain poorly understood, particularly in settings of moderate-to-high malaria transmission. We investigated associations between *Kelch13* and partner drug mutation prevalence and monthly confirmed malaria incidence across 16 Ugandan districts from 2017 to 2022, using integrated molecular, epidemiological, climatic, malaria-control, and antimalarial resistance data and negative binomial regression. Combined prevalence of WHO-validated and candidate *Kelch13* mutations increased over the study period. Each 10-percentage-point increase in *Kelch13* prevalence was associated with a 13.1% higher incidence after multivariable adjustment (aIRR 1.131, 95% CI 1.090–1.174). Furthermore, each 10% increase in *pfcrt* K76T was associated with a 24% increase in malaria transmission. As expected, ITN distribution ≥80%, IRS coverage ≥90%, and higher altitude were associated with lower incidence, whereas temperatures of 25–30°C, relative humidity of 60–80%, and higher precipitation were associated with higher incidence. The persistence of the *Kelch13* and *pfcrt* K76T association after adjustment suggests that expanding ART-R-associated parasite populations are independently associated with malaria incidence. The relationship may be bidirectional. These findings support interpreting ART-R within its ecological and intervention context and integrating molecular, epidemiological, and malaria-control surveillance to safeguard antimalarial efficacy.

## 1. Introduction

Antimalarial drug resistance is the cumulative product of a continuing evolutionary contest between parasite adaptation and efforts to maintain effective malaria treatment (1, 2). Antimalarial drugs remain essential for reducing malaria morbidity, mortality and onward transmission, but repeated drug exposure exerts strong selective pressure on *Plasmodium* populations, favouring resistance-associated variants that survive treatment and subsequently increase in frequency (3, 4). As these variants emerge, expand, and become established, treatment strategies and public health policy respond and adapt to preserve therapeutic efficacy and limit further spread. Molecular surveillance provides a means to detect and monitor these changes by observing the frequency and geographic distribution of resistance-associated parasite genotypes over time (5).

Monitoring the emergence and spread of artemisinin partial resistance (ART-R) is of particular importance as artemisinin-based combination therapies (ACTs) remain the foundation of treatment for uncomplicated and severe *Plasmodium falciparum* malaria across sub-Saharan Africa (6, 7). Resistance to the fast-acting component of ACT, artemisinin, is emerging and spreading in malaria-endemic regions across Africa and has become established in several African malaria-endemic countries (8). Genetic polymorphisms in the propeller domain of the *P. falciparum Kelch13* gene are clinically and experimentally associated with ART-R, and the strength of evidence linking individual mutations to ART-R is characterised and listed in the World Health Organization (WHO) compendium of molecular markers for antimalarial drug resistance (9). Distinct African- and region-specific *Kelch13* variants have emerged, including C469Y, R561H, A675V and P441L (10–13), associated with ART-R phenotypes, including delayed parasite clearance, in experimental or clinical studies. Five African malaria-endemic countries were designated as having WHO-confirmed ART-R: Eritrea, Rwanda, Uganda, the United Republic of Tanzania, and, most recently, Ethiopia. Additional surveillance and clinical reports suggest ART-R may be emerging in Namibia, Sudan and Zambia (14–16).

Despite advances in understanding the emergence, spread and clinical significance of *Kelch13* mutations, their contribution to malaria transmission and population-level disease burden remains poorly understood. These associations are of importance in sub-Saharan Africa, where malaria transmission is often higher, infections are more frequently polyclonal, population immunity is greater, and malaria-control intervention coverage is more variable than in South East Asia, where ART-R first emerged (1, 2). At the population level, changes in malaria incidence may simultaneously reflect climatic variation, vector abundance, population immunity, prompt access to effective treatment, chemoprevention coverage, and the introduction, intensification or withdrawal of vector-control interventions such as insecticide-treated nets (ITNs) and indoor residual spraying (IRS) (17, 18). The emergence of resistance to ACT partner drugs and other antimalarials may further alter these dynamics by increasing the risk of treatment failure. Determining whether the expansion of *Kelch13* mutant parasite populations is independently associated with malaria incidence, after accounting for these competing influences, is therefore important to understand the broader public health consequences of ART-R in high-burden African settings. Uganda provides a particularly relevant setting for examining these relationships, given its high malaria burden and the rapidly increasing prevalence of multiple *Kelch13* ART-R-associated mutations between 2017 and 2022 (19).

We investigated whether the increasing prevalence of *Kelch13* mutations was associated with malaria incidence across 16 malaria-endemic districts in Uganda during the study period. Longitudinal parasite genetic surveillance data were integrated with malaria incidence, climatic, and malaria intervention data to evaluate this association. We accounted for major determinants of malaria incidence, including temperature, relative humidity, precipitation, altitude, ITN distribution, IRS, chemoprevention in pregnant women, broader antimalarial drug resistance patterns, and malaria service disruptions associated with the COVID-19 pandemic.

## 2. Materials and Methods

### 2.1. Data collection

We used routinely collected malaria epidemiological, molecular surveillance, climate, malaria-control, and COVID-19 data to investigate malaria transmission in Uganda between 1 January 2017 and 31 December 2022. Data on the prevalence of *Kelch13* ART-R-associated mutations were obtained from previously published molecular surveillance data (13) and accessed through the WWARN Artemisinin Molecular Surveyor (www.iddo.org/wwarn/tracking-resistance/artemisinin-molecular-surveyor; accessed 26 June 2025). The mutations included in the analysis were WHO-validated ART-R markers A675V, R561H, and C469Y and the WHO-candidate markers C469F and P441L (9). The annual mutation prevalence was retrieved for each study district between 2017 and 2022.

To account for resistance-associated genotypes affecting other antimalarials used in Uganda, we included data of previously published molecular surveillance from the WWARN ACT Partner Drug Surveyor (www.iddo.org/wwarn/tools-surveyors/act-partner-drug-molecular-surveyor; accessed 1 July 2026) and the WWARN SP Surveyor (https://www.iddo.org/wwarn/tools-surveyors/sp-molecular-surveyor; accessed 1 July 2026), with a focus on the markers *pfcrt* K76T; *pfmdr1* N86Y, Y184F and D1246Y; *pfdhfr* N51I, C59R, and S108N; and *pfdhps* A437G, K540E and A581G. The prevalence estimates were obtained from existing geospatial models of the markers *pfcrt* K76T (20) *pfmdr1* N86Y, Y184F, and D1246Y; and *dhps* A437G, K540E, and A581G (20, 21). These markers are clinically and experimentally linked with altered susceptibility to ACT partner drugs (*pfcrt*/*pfmdr1*) and sulfadoxine-pyrimethamine (SP) resistance (*dhfr*/*dhps*), relevant to intermittent preventive treatment in pregnancy (IPTp) (**Table S1**), and were evaluated as candidate explanatory variables in the regression analyses (9).

The primary analyses included 16 districts for which corresponding molecular surveillance and malaria incidence data were available: Agago, Amolatar, Arua, Hoima, Jinja, Kaabong, Rukiga, Kanungu, Kapchorwa, Kasese, Katakwi, Koboko, Kole, Lamwo, Mubende, and Tororo. Routine malaria epidemiological data were obtained from Uganda’s Health Management Information System through the District Health Information Software 2 (DHIS2) platform (https://dhis2.org/metadata-package-downloads/; accessed on 1 July 2023). Monthly data from January 2017 to December 2022 were extracted for confirmed malaria cases. Confirmed malaria cases were defined as the sum of malaria diagnoses confirmed by blood-smear microscopy and rapid diagnostic testing. District population estimates were obtained from the Uganda Bureau of Statistics (https://www.ubos.org/explore-statistics/0/; accessed 6 June 2025) and were used as denominators to calculate monthly malaria incidence, expressed as confirmed malaria cases per 1,000 population.

For descriptive analyses, malaria epidemiological indicators were evaluated across the 136 Ugandan districts as defined by the 2023 administrative structure. These included confirmed malaria cases, malaria-associated deaths, hospitalisations, and outpatient consultations and were standardised by district population to facilitate geographical and temporal comparisons. To account for potential disruption to health services and malaria control during the COVID-19 pandemic, epidemiological data on COVID-19 in Uganda were obtained for the period 1 January 2020 to 31 December 2022. Estimated new confirmed COVID-19 cases were obtained from the COVID-19 Data Repository of the Johns Hopkins University Centre for Systems Science and Engineering and accessed through Our World in Data (OWID) (22).

Monthly climate data for the study period were obtained from the World Bank Climate Change Knowledge Portal (www.climateknowledgeportal.worldbank.org; accessed 6 June 2025). Variables included precipitation, relative humidity, and temperature. District altitude was also included as a geographical covariate (accessed via https://elevation.city/ug/4sklr#google_vignette; 1 July 2023). Data on malaria vector-control interventions were obtained from the Uganda Ministry of Health National Malaria Elimination Programme through DHIS2. Annual ITN distribution relative to the district population and annual IRS coverage were also collected for the districts of interest during the same period. The collected data on the ITN coverage were classified as ≥80% or <80% (23), while annual IRS coverage was classified as ≥90% or <90% (including areas without IRS). The district-level uptake estimates for intermittent preventive treatment in pregnancy with at least three doses of sulfadoxine-pyrimethamine (IPTp-SP3) were obtained from published district maps for Uganda for 2017–2022 and categorised as 0–33%, 34–65%, and 66–98% (24).

### 2.2. Data Management and Statistical Analysis

Epidemiological, molecular surveillance, climate, malaria-control, and COVID-19 data were harmonised into a district-month analytical dataset. Monthly malaria incidence, defined as confirmed malaria cases per 1,000 population, was the primary outcome. The primary exposure was the combined prevalence of the five WHO-validated and candidate *Kelch13* ART-R mutations considered in this study. For each district-year, the annual *Kelch13* prevalence estimate was assigned to each monthly malaria-incidence observation within the corresponding district and calendar year. Thus, *Kelch13* prevalence varied by district and year, while malaria incidence varied by district and month.

Associations between combined *Kelch13* mutation prevalence and malaria incidence were estimated using negative binomial regression in Stata version 18 (StataCorp LLC, College Station, Texas, USA), accounting for overdispersion in the outcome. Each pre-defined explanatory variable was first assessed in bivariate analysis, and variables associated with malaria incidence at *p*<0.20 were considered for multivariable modelling. A forward selection approach was used to construct the final model, with variables retained where they improved model fit. Random effects for district and year were included to account for clustering of repeated observations. Multicollinearity was assessed using variance inflation factors (VIFs). When strong correlations between predictors were identified, pairwise correlations, AIC, and the established relevance of each marker to antimalarial drug susceptibility and current treatment use in Uganda were used to guide variable retention.

### 2.3. Data Visualisation and Spatial-Temporal Mapping

Spatial and temporal patterns in malaria incidence and *Kelch13* mutation prevalence were visualised using district-level maps and graphical summaries. Choropleth maps of Uganda were generated to illustrate the geographical distribution of combined *Kelch13* mutation prevalence categories across the 16 study districts, while district-specific panel plots displayed annual combined and mutation-specific *Kelch13* prevalence alongside monthly malaria incidence. Additional district-level maps illustrated annual malaria incidence, malaria-associated mortality, and hospitalisation rates between 2017 and 2022. District-level data were linked to Uganda administrative boundaries obtained from the GeoBoundaries open-access database (25) and visualised using GeoPandas and Plotly in Python 3.9. Boundary geometries were projected to WGS84 (EPSG:4326), and the code used to generate the spatial visualisations is available at www.tinyurl.com/vanwyk-k13-2026.

### 2.4. Ethics Statement

This study was approved by the University of Cape Town, Faculty of Health Sciences Human Research Ethics Committee (HREC REF: 885/2025). Permission to conduct the study and to access *P. falciparum* genotypes, IRS, and ITN data in relevant Ugandan districts was granted by the Ugandan Ministry of Health’s National Malaria Elimination Division and the Infectious Diseases Research Collaboration. All data used were aggregate data with no risk of patient identification. Confidentiality of information was maintained by restricting access to study data to the principal investigator or designated study staff.

## 3. Results

### 3.1. Malaria Incidence and Control Interventions

All districts in Uganda were malaria-endemic; however, the malaria burden exhibited spatial and temporal heterogeneity across Uganda between 2017 and 2022 (**Figure 1**). The malaria incidence was consistently highest in several northern districts, although both its magnitude and geographical distribution varied between years. Similar spatial and temporal heterogeneity was observed for malaria-associated mortality and outpatient malaria burden. Across the 16 districts investigated (**Figure S1**), the dataset comprised monthly observations from January 2017 to December 2022, totalling 1,152 district-month observations. Median monthly malaria incidence was 263 cases per 1,000 population (IQR 137–524; range 14– 1,734), with the highest median incidence observed in Lamwo (1,330 cases per 1,000; range 322–1,734) and the lowest in Rukiga (31 cases per 1,000; range 14–72). Vector control coverage varied across districts and years; ITN distribution reached ≥80% of the district population in 44.8% of district-years, while IRS coverage ≥90% occurred in 16.7% of district-years (**Table 1**; **Figure 2**).

**Figure 1.**
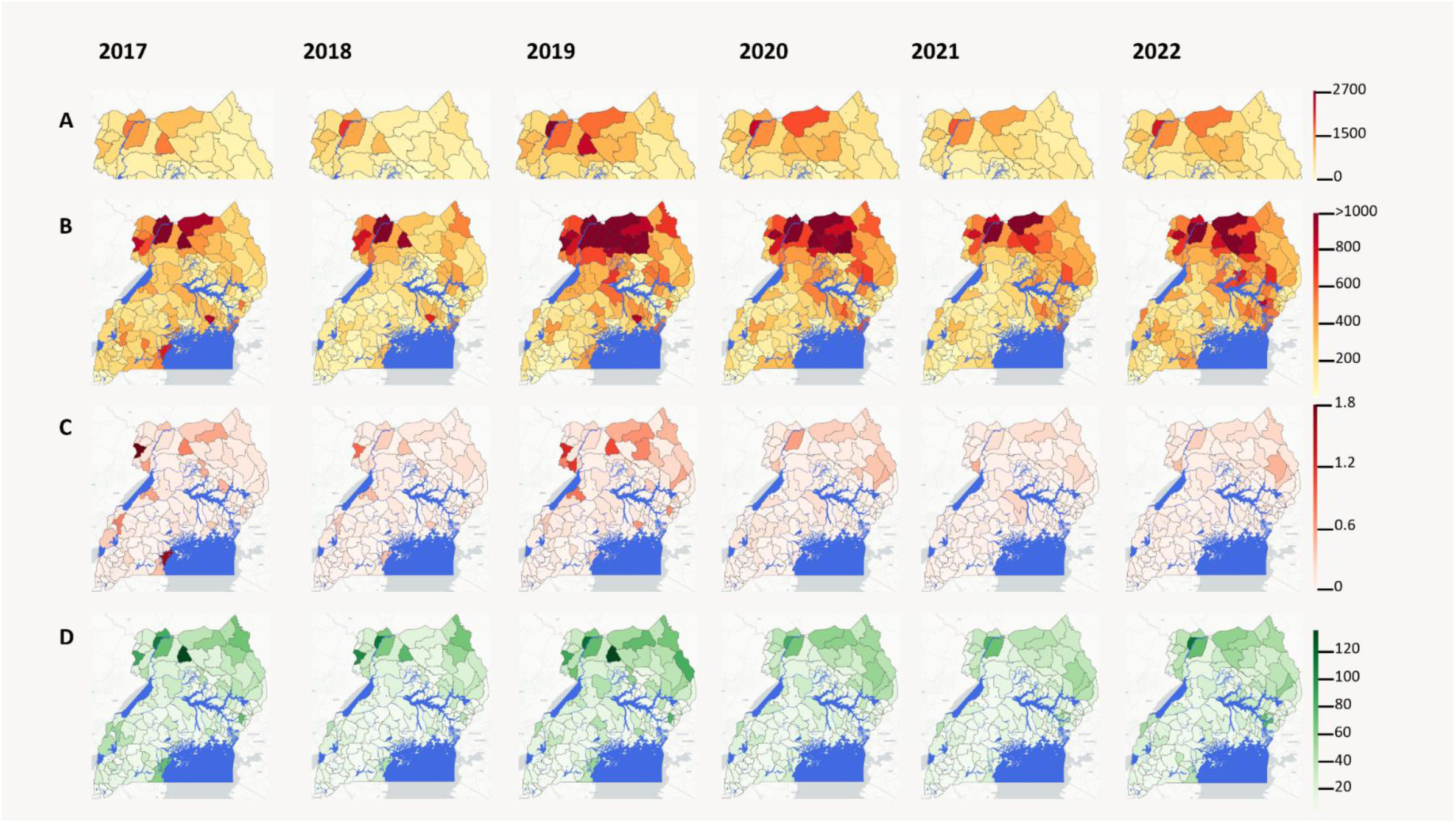
Spatial and temporal distribution of malaria burden in Uganda, 2017–2022. Annual district-level malaria indicators are shown from 2017 to 2022. (A) Malaria incidence among northern districts, displayed separately using an expanded scale to preserve variation among high-burden settings; (B) malaria incidence across Uganda; (C) malaria-associated mortality; and (D) malaria outpatient consultation rates. All indicators are expressed per 1,000 population. Colour intensity represents increasing incidence or rate within each panel.

**Figure 2.**
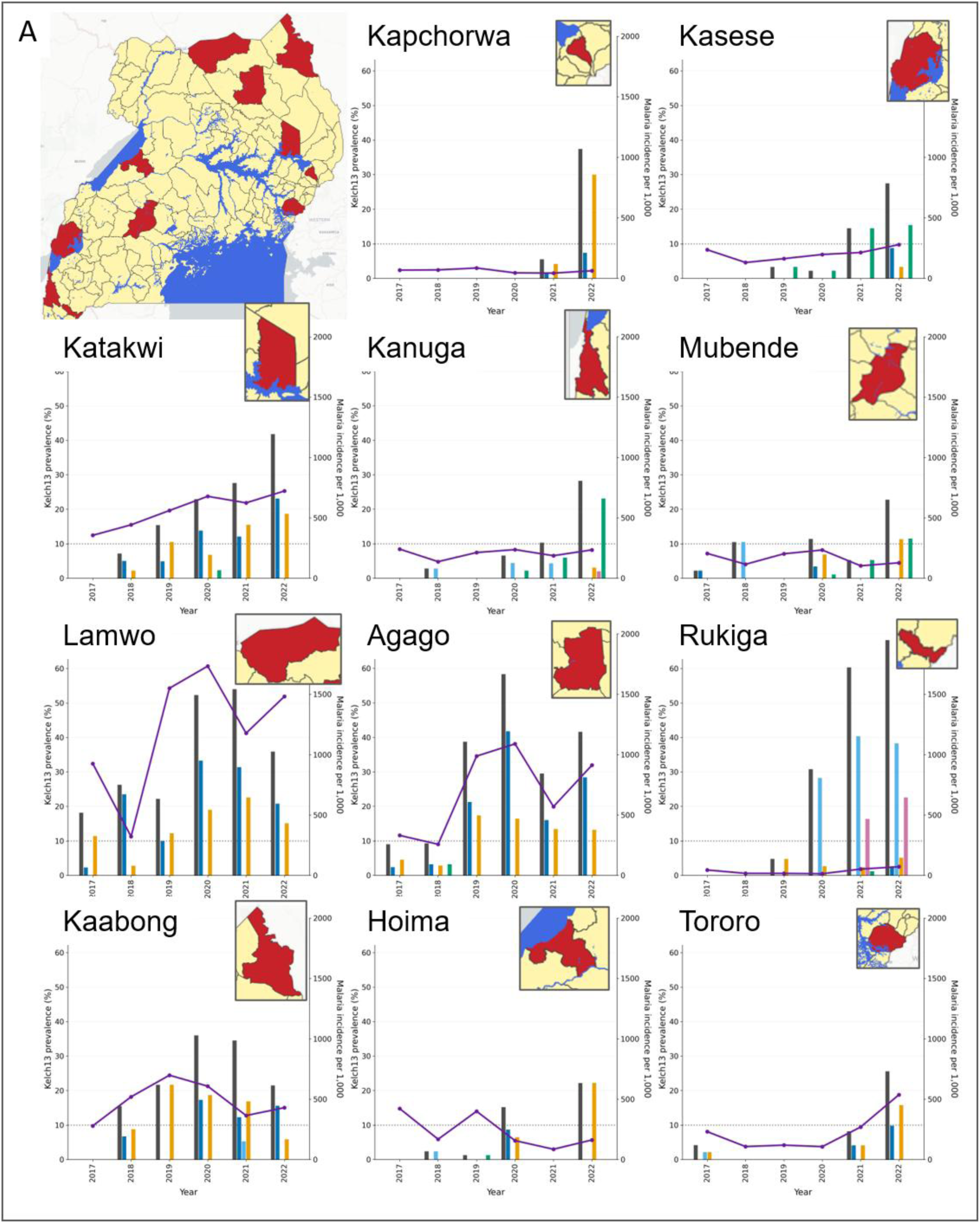
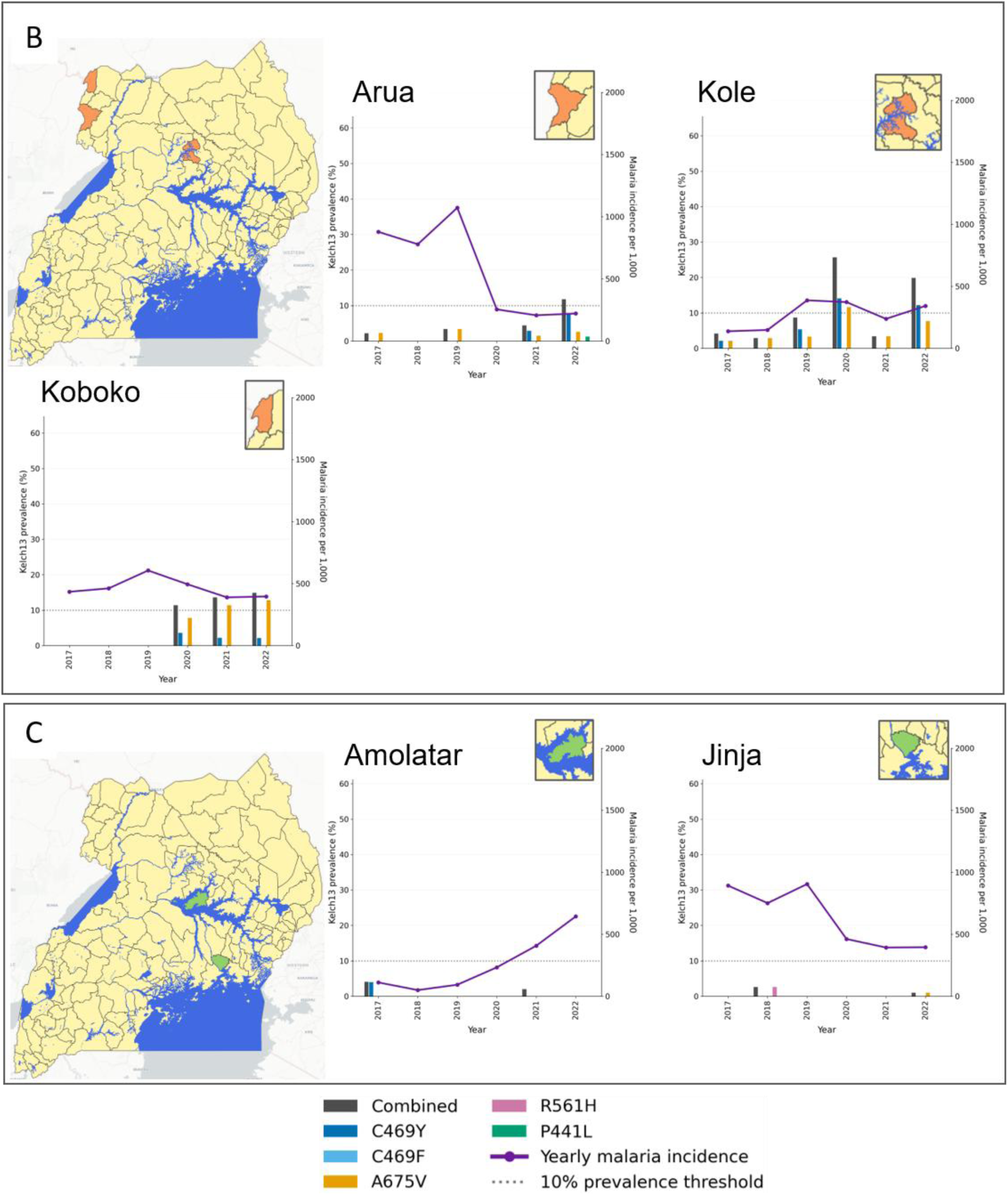
Spatial-temporal distribution of malaria incidence and *Kelch13* mutation prevalence across 16 districts in Uganda, 2017–2022. Districts were grouped according to the combined annual prevalence of validated and candidate Kelch13 resistance-associated mutations as follows: Figure 2.A) ≥20%; Figure 2.B) 10 to <20%; or Figure 2. C) <10%. For each district, the purple line represents malaria incidence (per 1000), and the coloured bars show the annual prevalence of individual *Kelch13* mutations (C469Y, C469F, A675V, R561H, and P441L), along with the combined prevalence. The dashed horizontal line denotes the WHO threshold of concern (10%). Insets indicate the geographical location of each district within Uganda.

**Table 1.** Descriptive characteristics of the 16 study districts investigated, Uganda, 2017–2022^1^.

| Characteristic | Summary |
| --- | --- |
| **Study characteristics** |  |
| Study districts, n | 16 |
| District-years, n | 96 |
| District-month observations, n | 1,152 |
| **Malaria epidemiology** |  |
| Median monthly malaria incidence, cases per 1,000 population | 263.2 (137.5–524.0); |
| Monthly range | 13.7–1,733.7 |
| **Kelch13 molecular surveillance** |  |
| Combined annual *Kelch13* mutation prevalence<sup>2</sup>, % | 4.6 (0–21.8) |
| District-year range, % | 0–68.2 |
| Combined *Kelch13* prevalence<sup>2</sup> <10% | 57/96 (59.4%) |
| Combined *Kelch13* prevalence<sup>2</sup> 10% to <20% | 13/96 (13.5%) |
| Combined *Kelch13* prevalence<sup>2</sup> ≥20% | 26/96 (27.1%) |
| **Climate and geography** |  |
| Median monthly temperature, mean °C | 27.5 (26.3–29.3); |
| Monthly range, °C | 21.7–35.4 |
| Monthly temperature 25–30°C | 810/1,152 (70.3%) |
| Median monthly relative humidity, % | 72.7 (64.2–77.5) |
| Monthly range, % | 27.8–86.9 |
| Monthly relative humidity 60–80% | 790/1,152 (68.6%) |
| Median [and mean] monthly precipitation, mm | 131.8 (65.5–197.9) |
| Monthly range, mm | 0–801.4 |
| District altitude, metres above sea level | 1,241 (1,093–1,363) |
| District range, metres above sea level | 1,000–1,915 |
| **Malaria-control interventions** |  |
| ITN distribution ≥80% of the district population | **43/96 (44.8%)** |
| IRS coverage ≥90% | 16/96 (16.7%) |
| IPTp-SP3 uptake 0–33% | 38/96 (39.6%) |
| IPTp-SP3 uptake 34–65% | 43/96 (44.8%) |
| IPTp-SP3 uptake 66–98% | 15/96 (15.6%) |
| **Additional molecular resistance marker** |  |
| *pfcr*t K76T predicted prevalence, %, median (IQR) | 14.8 (3.6–29.5) |
| District-year range, % | 0.9–70.3 |
| **COVID-19** |  |
| Confirmed monthly COVID-19 cases, per 1,000 population | 0 (0–0) |
| Monthly range | 0–366.2 |

<sup>1</sup>Summary statistics were calculated at the temporal resolution of each variable: malaria incidence, temperature, relative humidity, precipitation and COVID-19 incidence were summarised across 1,152 district-month observations; *Kelch13* prevalence, ITN distribution, IRS coverage, IPTp-SP3 uptake and predicted *pfcr*t K76T prevalence were summarised across 96 district-years; altitude was summarised across the 16 study districts.
<sup>2</sup>The combined prevalence included the prevalences of the WHO-validated ART-R markers A675V, R561H, and C469Y and the WHO-candidate markers C469F and P441L (9).

### 3.2. Distribution of *Kelch13* Mutations

The combined prevalence of WHO-validated and candidate *Kelch13* mutations varied across districts and over time, and increased progressively over the study period (**Table 2** and **Figure 2).** In 2017, 15 of 16 districts had a combined *Kelch13* prevalence per district below 10%, one district had a prevalence between 10% and 20%, and none had a prevalence ≥20%. By 2022, 11 of 16 districts had combined *Kelch13* prevalence ≥20%, three had prevalence between 10% and 20%, and only two remained below 10%. Across all districts and years, combined district-specific *Kelch13* prevalence ranged from 0% to 68.2%, with the highest prevalences concentrated in northern and western districts.

**Table 2:**
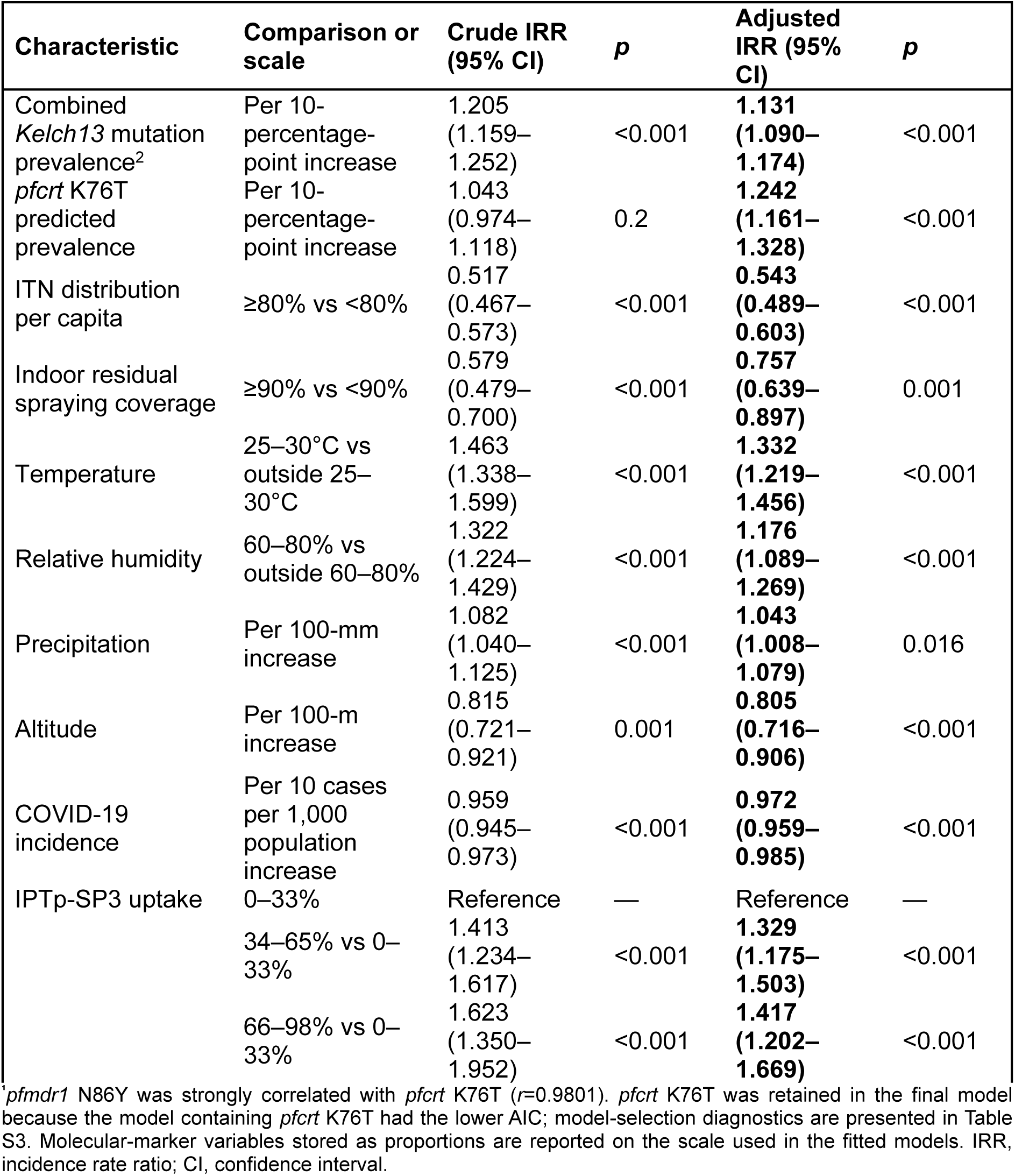
Association between *Kelch13* prevalence and malaria incidence in 16 Ugandan districts, 2017–2022^1^.

The individual ART-R-associated mutations showed distinct geographical patterns. C469Y and A675V were most prominent in northern Uganda bordering South Sudan, while P441L was predominantly concentrated in western districts bordering the Democratic Republic of the Congo. R561H and C469F were observed mainly in southwestern Uganda bordering Rwanda. In several districts, increases in combined *Kelch13* prevalence occurred during periods of high or increasing malaria incidence, although the timing and direction of these trends were not uniform across all sites (**Figure 2**).

### 3.3. *Kelch13* Mutation Prevalence and Malaria Incidence

Given the observed spatial and temporal heterogeneity, regression analyses were used to assess whether *Kelch13* prevalence was independently associated with malaria incidence after accounting for climatic, malaria-control, epidemiological and additional antimalarial resistance-related genetic markers (**Table 2** and **Tables S2** and **S3**). In univariate analysis, each 10-percentage-point increase in combined *Kelch13* prevalence was associated with a 20.5% increase in malaria incidence (cIRR 1.205, 95% CI 1.159–1.252; *p*<0.001). After multivariable adjustment, the association was attenuated but remained statistically significant, with each 10-percentage-point increase in *Kelch13* prevalence associated with a 13.1% increase in malaria incidence (aIRR 1.131, 95% CI 1.090–1.174; *p*<0.001) (**Table 2**).

Data on additional molecular markers of antimalarial resistance were limited, particularly for markers with incomplete district-year coverage and those not represented in the available surveillance datasets (**Table S1**). Among the available ACT partner-drug resistance-associated markers, *pfcrt* K76T was strongly correlated with *pfmdr1* N86Y prevalence (r=0.9801). To minimise multicollinearity, *pfcrt* K76T was retained in the final model based on improved model fit (**Table S3**). Each 10-percentage-point increase in predicted *pfcrt* K76T prevalence was associated with a 24.2% higher malaria incidence (aIRR 1.242, 95% CI 1.161–1.328; p<0.001).

Malaria-control interventions were independently associated with lower incidence. ITN distribution ≥80% was associated with a 45.7% lower malaria incidence (aIRR 0.543, 95% CI 0.489–0.603; p<0.001), while IRS coverage ≥90% was associated with a 24.3% lower incidence (aIRR 0.757, 95% CI 0.639–0.897; p=0.001). Temperatures of 25–30°C (aIRR 1.332, 95% CI 1.219–1.456; p<0.001) and relative humidity of 60–80% (aIRR 1.176, 95% CI 1.089–1.269; p<0.001) were associated with higher malaria incidence. A 100-mm increase in monthly precipitation was associated with a 4% higher malaria incidence (aIRR 1.043, 95% CI 1.008–1.079; *p*=0.016), whereas a 100m increase in altitude was associated with a 19.5% lower malaria incidence (aIRR 0.805, 95% CI 0.716–0.906; *p*<0.001).

A 10-case-per-1,000 population increase in COVID-19 incidence was associated with a 2.8% lower malaria incidence (aIRR 0.972, 95% CI 0.959–0.985; *p*<0.001). Compared with IPTp-SP3 uptake of 0–33%, an uptake of 34–65% (aIRR 1.329, 95% CI 1.175–1.503) and 66–98% (aIRR 1.417, 95% CI 1.202–1.669) was associated with higher malaria incidence (*p*<0.001 for both).

Model-based predictions from the final adjusted negative binomial model showed increasing predicted malaria incidence across the observed range of combined WHO-validated and candidate *Kelch13* prevalence, with apparent tapering for the two observations with very high *Kelch13* prevalence (>60%) (**Figure 3**).

**Figure 3.**
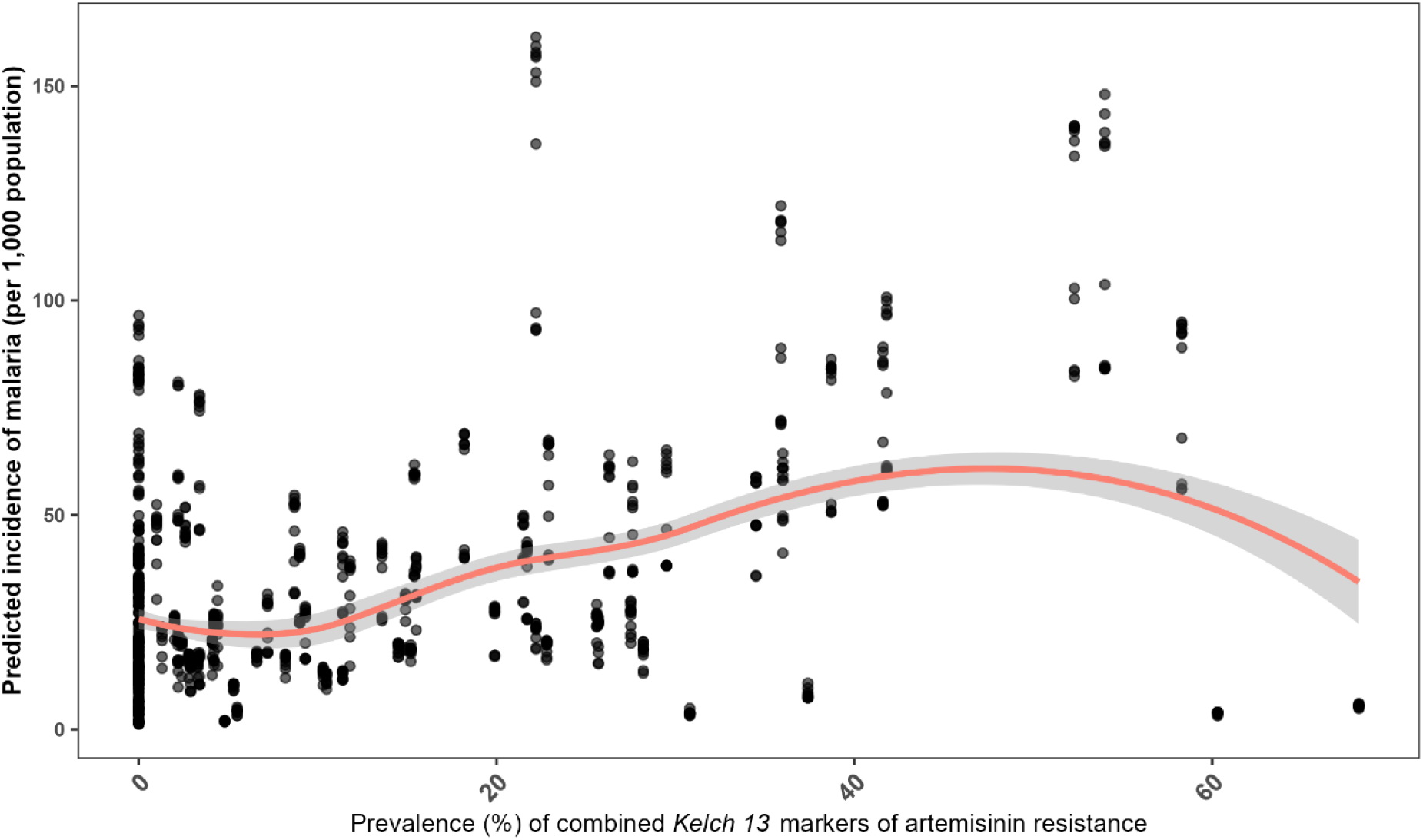
Predicted malaria incidence across the combined *Kelch13* mutation prevalence in the final adjusted model.

## 4. Discussion

The role of *Kelch13* mutants in driving ART-R is increasingly well characterised (26–28), as is the value of molecular surveillance for monitoring ART-R emergence and spread. However, their contribution to malaria transmission and population-level burden remains less well understood, particularly in areas of moderate-to-high malaria transmission intensity. In this study, each 10-percentage-point increase in combined *Kelch13* prevalence was associated with a 20.5% increase in malaria incidence in unadjusted analysis and a 13.1% increase after adjustment for climatic, vector-control, epidemiological and antimalarial resistance-related factors. The attenuation after adjustment indicates that part of the crude association reflects the broader transmission environment, while the persistence of the association in the final model suggests that the expansion of *Kelch13* prevalence was independently associated with increased malaria incidence at the population level. This association should nevertheless be interpreted within the broader complexity of malaria transmission, which is shaped not only by *Kelch13* mutations but by interacting parasite, treatment, health-system, ecological, and vector-control factors (17).

Uganda provides an important setting for investigating the population-level consequences of expanding *Kelch13* mutant parasite populations and the establishment of ART-R in high-burden African settings. Artemether-lumefantrine (AL) has been the first-line treatment for uncomplicated malaria since 2006, providing sustained population-level exposure to both artemisinin and lumefantrine (29). Clinical and experimental evidence of reduced artemisinin susceptibility was detected in northern Uganda between 2017 and 2019, followed by an increase in the prevalence of the *Kelch13* A675V and C469Y mutations, which were associated with prolonged parasite clearance and increased ring-stage survival (29, 30). Our data capture this period of expansion, during which combined *Kelch13* prevalence increased markedly between 2017 and 2022, with the highest prevalences occurring particularly in northern Uganda, where malaria transmission is high. By 2021–2022, *Kelch13* mutations exceeded 20% prevalence in 11 of 16 surveillance districts. Subsequent therapeutic efficacy studies in 2022–2023 provided clinical evidence of this evolving resistance landscape, with reduced artemether-lumefantrine efficacy in two of the three sites investigated, and C469Y- and A675V-carrying infections showed prolonged parasite clearance relative to wild-type infections at these study sites (31).

Of importance, *Kelch13* mutations do not contribute equally to the ART-R phenotype (9, 26, 27). WHO classifications reflect differing levels of clinical and experimental evidence linking individual variants to delayed parasite clearance, and validated, candidate and potential mutations differ in the magnitude and consistency of their phenotypic effects (9, 27, 28). This distinction is particularly relevant in Uganda, where resistance-associated mutations have emerged in different epidemiological and geographical settings, often reflecting cross-border population mobility (1). Transmission intensity also modifies the phenotypic expression and detection of ART-R. Parasite clearance is generally faster in moderate-to-high transmission settings, where acquired immunity and multiclonal infections can obscure delayed clearance despite the presence of resistance-associated genotypes (27, 30). In this study, we combined *Kelch13* mutations to capture the population-level expansion of resistance-associated parasite genotypes while recognising that individual mutations may differ substantially in their contribution to ART-R. These trends should therefore be interpreted within the context of long-standing reliance on AL, since repeated antimalarial exposure can favour resistant parasites that survive treatment, while changes in vector-control coverage may facilitate their expansion.

The association between increasing *Kelch13* prevalence and malaria incidence is biologically plausible and may operate bidirectionally. ART-R delays parasite clearance following treatment (27, 28, 31) allowing resistant parasites to persist for longer, increasing the residual parasite biomass that must be eliminated by the ACT partner drug, and thereby increasing selective drug pressure on the partner drug, lumefantrine and subsequently the risk of treatment failure. ART-R and delayed parasite clearance have also been associated with increased gametocyte carriage and greater potential for onward transmission (32). Conversely, high-burden settings generate larger parasite populations and more frequent infections, creating more opportunities for resistant parasites to emerge and spread. Although acquired immunity means that a greater proportion of infections may remain asymptomatic and untreated (33), the high absolute burden of symptomatic malaria can still result in extensive antimalarial use and selection pressure. Selective pressure may also extend to ACT partner drugs, such as lumefantrine, as reflected by increasing *pfcrt* K76T and *pfmdr1* N86Y prevalence wherever it is widely used (1). In this study, each 10-percentage-point increase in *pfcrt* K76T was associated with a 24% higher malaria incidence, and was strongly correlated with *pfmdr1* N86Y, illustrating the difficulty of separating correlated components of the broader antimalarial-resistance background. Together, these findings suggest that the relationship between ART-R and malaria burden is shaped by both parasite resistance and the surrounding transmission environment.

Indeed, the wider model results showed that *Kelch13* prevalence operates within a strong ecological and intervention context. ITN distribution ≥80% was associated with a 45.7% lower malaria incidence, while IRS coverage ≥90% was associated with a 24.3% lower incidence. Therefore, maintaining strong vector-control coverage in ART-R-threatened areas is essential. Higher temperature and relative humidity were associated with increased incidence, precipitation showed a smaller positive association, and increasing altitude was associated with lower incidence, all consistent with the well-established influence of environmental conditions on malaria transmission and burden. The inverse association between reported COVID-19 burden and recorded incidence is more plausibly explained by changes in health-seeking behaviour, diagnostic activity or routine reporting during the pandemic than by a protective effect on malaria transmission. The unexpected positive association between IPTp-SP3 uptake and malaria incidence may reflect underlying differences in malaria burden, service utilisation or surveillance completeness across districts rather than a direct effect of IPTp-SP3 itself. These ecological associations may therefore be subject to residual confounding despite multivariable adjustment.

These findings reinforce the importance of strengthening malaria control and surveillance in the face of rapidly increasing ART-R in high-burden areas. Interruption of vector control, diagnosis, treatment, or preventive services can increase the malaria burden and the number of infections requiring ACT treatment, both antimalarial selection pressure on parasite populations and treatment-seeking costs for households and health providers (7, 28). In settings where resistance-associated *Kelch13* mutations are already established, such conditions could favour the continued expansion and onward transmission of *Kelch13* genotypes. This concern is increasingly relevant in the current constrained-financing environment (6), where disruptions to surveillance, vector control, diagnostics, medicines, and other essential malaria services may weaken both malaria control and the capacity to detect changes in the antimalarial resistance landscape.

Several limitations to this study should be considered. First, this ecological analysis cannot establish a causal relationship between the increase in *Kelch13* prevalence and malaria incidence; the association may be bidirectional. Second, the combined *Kelch13* exposure incorporated mutations with differing levels of evidence for ART-R-related delayed parasite clearance and increased gametocytes and malaria transmission; these do not imply equivalent biological effects across variants. Surveillance should distinguish among individual *Kelch13* mutations and account for transmission setting, because both genotype-phenotype relationships and the detectability of delayed parasite clearance vary across epidemiological contexts (27). Third, annual molecular prevalence estimates were linked to monthly malaria observations, and residual confounding by unmeasured or imperfectly measured transmission-related factors may remain despite the inclusion of district- and year-level random effects and multiple climatic, intervention-related, and resistance-related covariates. Finally, malaria incidence was derived from routine health-facility data and may have been influenced by variation in healthcare access, diagnostic activity, reporting completeness and service disruptions, including those incurred during the COVID-19 pandemic. Despite these limitations, to our knowledge, this is the first study in Uganda to evaluate the population-level association between longitudinal prevalence of WHO-validated and candidate *Kelch13* ART-R markers and malaria incidence, while accounting for major climatic, malaria-control, epidemiological, and antimalarial resistance-related factors.

## 5. Conclusion

The study captures a period when resistance-associated *Kelch13* mutations expanded across Uganda as clinically confirmed ART-R was being established. It demonstrates that ART-R expansion was associated with a significant increase in malaria incidence even after accounting for major features of the transmission environment. Strengthening the completeness and continuity of molecular and clinical surveillance, alongside intensifying routine malaria-control services and specific antimalarial drug-resistance response strategies, including ACT diversification, will be essential for preventing the expansion of ART-R and the associated increase in malaria burden in Uganda and elsewhere in sub-Saharan Africa.

## Author Contributions

Conceptualization, S.v.W. and K.I.B.; methodology, S.v.W., K.I.B. and L.M.; investigation, S.v.W., L.M. K.I.B.; formal analysis, S.v.W., L.M., and K.I.B.; data curation, S.v.W., L.E.H. and J.G.; visualization, S.v.W.; supervision, K.I.B. and J.N.; project administration, S.v.W.; funding acquisition, K.I.B. and M.R.K.; writing—original draft preparation, S.v.W.; writing—review and editing, S.v.W., J.G., L.M., V.A., L.E.H., M.R.K., J.N. and K.I.B. All authors have read and agreed to the published version of the manuscript.

## Institutional Review Board Statement

The study was approved by the University of Cape Town Faculty of Health Sciences Human Research Ethics Committee (HREC REF: 885/2025).

## Data Availability Statement

The data supporting the findings of this study are provided in the Supplementary Materials and are available from the publicly accessible sources described in the manuscript. *Kelch13* molecular surveillance data were obtained from the WWARN Artemisinin Molecular Surveyor (https://www.iddo.org/wwarn/tracking-resistance/artemisinin-molecular-surveyor), while additional antimalarial resistance-marker data were obtained from the WWARN ACT Partner Drug Surveyor (https://www.iddo.org/wwarn/tools-surveyors/act-partner-drug-molecular-surveyor) and WWARN SP Surveyor (https://www.iddo.org/wwarn/tools-surveyors/sp-molecular-surveyor). Analysis and figure-generation code are available at https://tinyurl.com/vanwyk-k13-2026.

## Abbreviations

ACT: artemisinin-based combination therapy
AIC: Akaike information criterion
aIRR: adjusted incidence rate ratio
AL: artemether-lumefantrine
ART-R: artemisinin partial resistance
CI: confidence interval
cIRR: crude incidence rate ratio
COVID-19: coronavirus disease 2019
DHIS2: District Health Information Software 2
EDCTP3: European and Developing Countries Clinical Trials Partnership 3
EPSG: European Petroleum Survey Group
HREC: Human Research Ethics Committee
IDRC: Infectious Diseases Research Collaboration
IPTp: intermittent preventive treatment in pregnancy
IPTp-SP3: intermittent preventive treatment in pregnancy with at least three doses of sulfadoxine-pyrimethamine
IQR: interquartile range
IRR: incidence rate ratio
IRS: indoor residual spraying
ITN: insecticide-treated net
MARC SE-Africa: Mitigating Antimalarial Resistance Consortium in South-East Africa
OWID: Our World in Data
P. falciparum: Plasmodium falciparum
pfcrt: Plasmodium falciparum chloroquine resistance transporter gene
pfdhfr: Plasmodium falciparum dihydrofolate reductase gene
pfdhps: Plasmodium falciparum dihydropteroate synthase gene
pfmdr1: Plasmodium falciparum multidrug resistance 1 gene
SD: standard deviation
SP: sulfadoxine-pyrimethamine
UCT: University of Cape Town
VIF: variance inflation factor
WGS84: World Geodetic System 1984
WHO: World Health Organization
WWARN: Worldwide Antimalarial Resistance Network

