## Appendix for "Expanding *Kelch13* artemisinin resistance markers are associated with higher malaria incidence in Uganda"

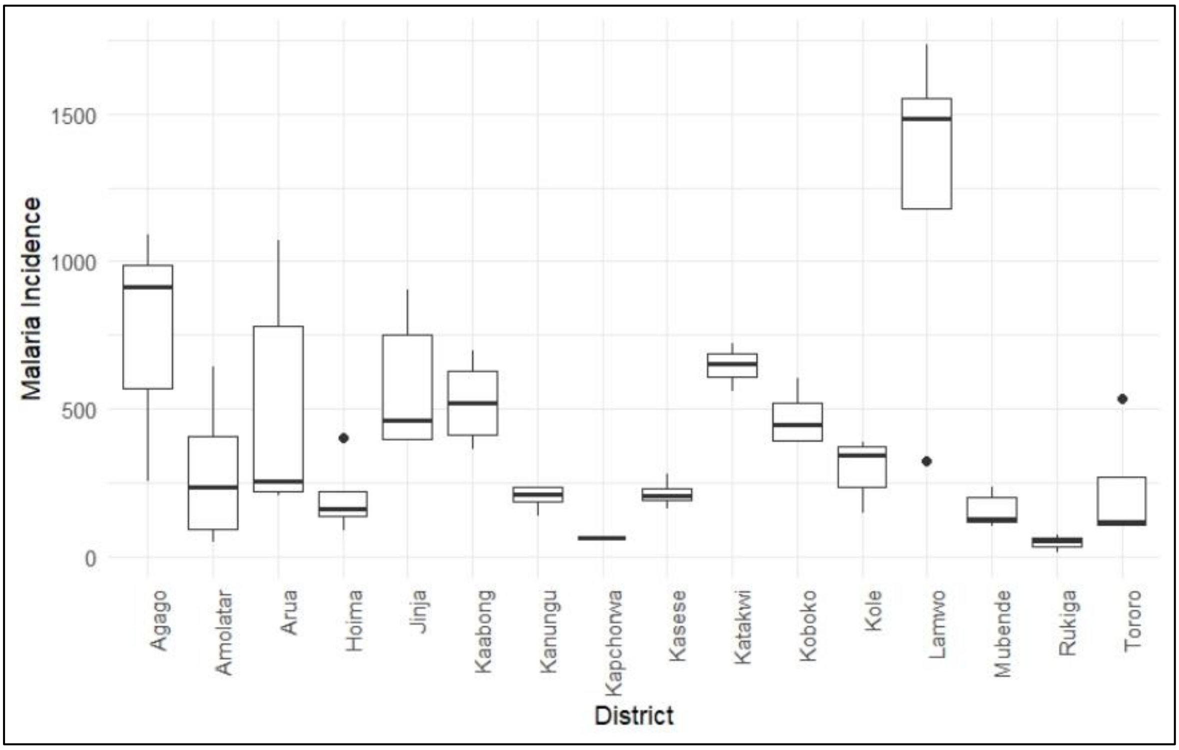


**Supplementary Figure 1:** Distribution of the monthly malaria incidence per 1000 population in the 16 study districts between 2017 and 2022.

**Supplementary Tables:**
**Table S1. Prevalence of markers of partner drug and sulfadoxine-pyrimethamine resistance[1]**

| District | Year | *pfcrt* K76T (%) | *pfmdr1* N86Y (%) | *pfmdr1* Y184F (%) | *pfmdr1* D1246Y (%) | *pfdhfr* S108N (%) | *pfdhfr* N51I (%) | *pfdhfr* C59R (%) | *pfdhps* A437G (%) | *pfdhps* K540E (%) | *pfdhps* A581G (%) |
|---|---|---|---|---|---|---|---|---|---|---|---|
| Agago | 2017 | 9,2 | 13,3 | 14,9 | 16,6 | 100,0 | 100,0 | 90,5 | 95,5 | 93,0 | 9,5 |
| Agago | 2018 | 29,1 | 23,1 | 18,9 | 15,6 | 100,0 | 100,0 | 96,6 | 95,8 | 90,0 | 8,8 |
| Agago | 2019 | 11,8 | 3,3 | 2,4 | 1,7 | 100,0 | 100,0 | 100,0 | 95,2 | 89,4 | 6,1 |
| Agago | 2020 | 1,2 | 1,0 | 66,4 | 67,2 | | | | 96,0 | 91,5 | 6,1 |
| Agago | 2021 | 68,8 | 69,2 | 69,8 | 41,1 | 100,0 | 100,0 | 46,5 | | | |
| Agago | 2022 | 26,8 | 19,3 | 13,9 | 11,5 | 100,0 | 100,0 | 90,7 | | | |
| Amolatar | 2017 | 4,6 | 6,5 | 7,9 | 9,7 | 100,0 | 100,0 | 94,9 | 96,8 | 95,6 | 11,6 |
| Amolatar | 2018 | 15,4 | 11,3 | 8,6 | 6,7 | 100,0 | 100,0 | 100,0 | 97,2 | 93,8 | 10,9 |
| Amolatar | 2019 | 4,9 | 3,6 | 2,5 | 1,8 | 100,0 | 100,0 | 100,0 | 96,6 | 92,6 | 7,4 |
| Amolatar | 2020 | 1,1 | 1,0 | 63,5 | 64,8 | | | | 97,3 | 94,2 | 7,2 |
| Amolatar | 2021 | 67,2 | 67,9 | 68,5 | 49,8 | | | | | | |
| Amolatar | 2022 | 30,8 | 21,5 | 15,0 | 12,0 | | | | | | |
| Arua | 2017 | 3,6 | 5,4 | 7,4 | 9,8 | 100,0 | 100,0 | 84,1 | 93,8 | 90,1 | 18,0 |
| Arua | 2018 | 32,8 | 27,2 | 23,1 | 19,6 | 100,0 | 100,0 | 90,5 | 94,0 | 85,7 | 16,1 |
| Arua | 2019 | 15,4 | 5,0 | 3,6 | 2,5 | 100,0 | 100,0 | 93,8 | 94,4 | 84,2 | 10,8 |
| Arua | 2020 | 1,6 | 1,3 | 63,3 | 64,5 | | | | 94,5 | 88,2 | 10,6 |
| Arua | 2021 | 67,0 | 67,9 | 69,0 | 29,7 | | | | | | |
| Arua | 2022 | 20,5 | 15,6 | 12,0 | 10,7 | | | | | | |
| Hoima | 2017 | 3,0 | 4,2 | 5,6 | 7,5 | | | | 95,9 | 94,6 | 17,0 |
| Hoima | 2018 | 26,9 | 21,2 | 17,0 | 13,9 | 100,0 | 100,0 | 95,3 | 96,8 | 91,7 | 15,3 |
| Hoima | 2019 | 10,1 | 3,8 | 2,7 | 1,9 | 100,0 | 100,0 | 91,9 | 95,9 | 91,7 | 12,1 |
| Hoima | 2020 | 1,2 | 1,0 | 60,8 | 62,1 | | | | 96,4 | 93,2 | 9,8 |
| Hoima | 2021 | 64,7 | 65,8 | 67,0 | 44,6 | | | | | | |
| Hoima | 2022 | 28,3 | 20,4 | 14,7 | 11,9 | | | | | | |
| Jinja | 2017 | 2,5 | 3,4 | 4,3 | 5,3 | 100,0 | 100,0 | 100,0 | 96,6 | 95,9 | 11,1 |
| Jinja | 2018 | 12,3 | 8,5 | 6,3 | 4,7 | 100,0 | 100,0 | 97,1 | 96,9 | 94,4 | 9,1 |
| Jinja | 2019 | 3,3 | 2,8 | 2,0 | 1,4 | 100,0 | 100,0 | 100,0 | 96,5 | 93,1 | 7,4 |

| District | Year | *pfcrt* K76T (%) | *pfmdr1* N86Y (%) | *pfmdr1* Y184F (%) | *pfmdr1* D1246Y (%) | *pfdhfr* S108N (%) | *pfdhfr* N51I (%) | *pfdhfr* C59R (%) | *pfdhps* A437G (%) | *pfdhps* K540E (%) | *pfdhps* A581G (%) |
|---|---|---|---|---|---|---|---|---|---|---|---|
| Jinja | 2020 | 0,9 | 0,8 | 63,0 | 64,7 | | | | 97,2 | 95,3 | 6,1 |
| Jinja | 2021 | 68,0 | 68,9 | 69,4 | 55,3 | | | | | | |
| Jinja | 2022 | 32,5 | 22,9 | 15,3 | 11,6 | | | | | | |
| Kaabong | 2017 | 7,8 | 10,2 | 11,5 | 13,3 | | | | 94,2 | 91,2 | 7,7 |
| Kaabong | 2018 | 48,4 | 41,5 | 35,9 | 31,0 | 100,0 | 100,0 | 86,2 | 94,1 | 87,7 | 7,2 |
| Kaabong | 2019 | 25,3 | 3,4 | 2,5 | 1,9 | 100,0 | 100,0 | 93,3 | 94,2 | 86,9 | 5,4 |
| Kaabong | 2020 | 1,3 | 1,1 | 68,3 | 68,9 | | | | 94,9 | 89,0 | 5,1 |
| Kaabong | 2021 | 70,0 | 70,3 | 70,9 | 31,3 | | | | | | |
| Kaabong | 2022 | 19,0 | 13,4 | 9,9 | 8,5 | | | | | | |
| Kanungu | 2017 | 4,7 | 5,9 | 7,0 | 8,5 | 100,0 | 100,0 | 97,8 | 95,0 | 95,3 | 38,2 |
| Kanungu | 2018 | 21,5 | 16,9 | 13,5 | 11,1 | 100,0 | 100,0 | 97,2 | 94,3 | 93,1 | 32,4 |
| Kanungu | 2019 | 8,1 | 3,6 | 2,7 | 2,0 | 100,0 | 100,0 | 100,0 | 95,2 | 91,8 | 30,3 |
| Kanungu | 2020 | 1,3 | 1,2 | 53,2 | 54,4 | | | | 94,7 | 93,8 | 22,9 |
| Kanungu | 2021 | 56,8 | 58,0 | 59,2 | 27,7 | | | | | | |
| Kanungu | 2022 | 17,6 | 13,0 | 9,6 | 7,9 | | | | | | |
| Kapchorwa | 2017 | 3,0 | 4,7 | 6,2 | 8,1 | | | | 96,9 | 94,9 | 5,2 |
| Kapchorwa | 2018 | 13,9 | 9,7 | 7,1 | 5,3 | | | | 96,1 | 93,1 | 4,3 |
| Kapchorwa | 2019 | 3,7 | 5,5 | 3,6 | 2,4 | 100,0 | 100,0 | 100,0 | 96,6 | 92,6 | 3,8 |
| Kapchorwa | 2020 | 1,4 | 1,1 | 57,7 | 59,1 | | | | 97,1 | 93,9 | 3,7 |
| Kapchorwa | 2021 | 61,5 | 62,3 | 63,5 | 32,2 | | | | | | |
| Kapchorwa | 2022 | 20,7 | 15,2 | 11,4 | 9,5 | | | | | | |
| Kasese | 2017 | 4,5 | 6,0 | 7,5 | 9,2 | | | | 96,0 | 95,3 | 27,1 |
| Kasese | 2018 | 24,6 | 19,6 | 15,8 | 13,0 | 100,0 | 100,0 | 100,0 | 96,2 | 93,3 | 23,4 |
| Kasese | 2019 | 9,5 | 3,8 | 2,8 | 2,0 | 100,0 | 100,0 | 91,3 | 95,8 | 92,6 | 20,4 |
| Kasese | 2020 | 1,3 | 1,2 | 55,1 | 56,5 | | | | 96,0 | 94,3 | 17,0 |
| Kasese | 2021 | 59,2 | 60,3 | 61,4 | 33,2 | | | | | | |
| Kasese | 2022 | 21,6 | 15,8 | 11,5 | 9,2 | | | | | | |
| Katakwi | 2017 | 7,7 | 11,5 | 13,8 | 15,7 | | | | 96,9 | 95,0 | 7,2 |
| Katakwi | 2018 | 14,1 | 10,0 | 7,6 | 5,9 | 100,0 | 100,0 | 97,5 | 96,5 | 92,4 | 6,1 |

| District | Year | *pfcrt* K76T (%) | *pfmdr1* N86Y (%) | *pfmdr1* Y184F (%) | *pfmdr1* D1246Y (%) | *pfdhfr* S108N (%) | *pfdhfr* N51I (%) | *pfdhfr* C59R (%) | *pfdhps* A437G (%) | *pfdhps* K540E (%) | *pfdhps* A581G (%) |
|---|---|---|---|---|---|---|---|---|---|---|---|
| Katakwi | 2019 | 4,3 | 3,6 | 2,5 | 1,7 | 100,0 | 100,0 | 100,0 | 96,5 | 92,2 | 5,0 |
| Katakwi | 2020 | 1,1 | 0,9 | 61,4 | 62,1 | | | | 97,2 | 93,6 | 4,8 |
| Katakwi | 2021 | 63,6 | 63,6 | 64,3 | 44,8 | | | | | | |
| Katakwi | 2022 | 28,4 | 20,4 | 14,7 | 12,1 | | | | | | |
| Koboko | 2017 | 3,6 | 5,3 | 7,2 | 9,6 | | | | 92,1 | 86,8 | 15,8 |
| Koboko | 2018 | 35,7 | 30,1 | 25,8 | 22,3 | 100,0 | 100,0 | 91,3 | 91,9 | 81,6 | 15,2 |
| Koboko | 2019 | 17,9 | 5,4 | 3,9 | 2,8 | 100,0 | 100,0 | 100,0 | 92,5 | 79,0 | 10,4 |
| Koboko | 2020 | 1,8 | 1,5 | 64,2 | 65,2 | | | | 93,0 | 84,7 | 10,1 |
| Koboko | 2021 | 67,6 | 68,4 | 69,5 | 25,9 | | | | | | |
| Koboko | 2022 | 18,0 | 14,0 | 11,0 | 9,8 | | | | | | |
| Kole | 2017 | 7,9 | 11,9 | 13,8 | 16,0 | 100,0 | 100,0 | 97,6 | 96,3 | 94,5 | 11,0 |
| Kole | 2018 | 20,7 | 15,7 | 12,5 | 10,1 | 100,0 | 100,0 | 93,1 | 96,7 | 92,5 | 10,8 |
| Kole | 2019 | 7,5 | 3,2 | 2,3 | 1,6 | 100,0 | 100,0 | 92,0 | 96,0 | 91,6 | 7,1 |
| Kole | 2020 | 1,1 | 0,9 | 65,6 | 66,6 | | | | 96,6 | 93,5 | 7,2 |
| Kole | 2021 | 68,4 | 68,8 | 69,3 | 48,8 | | | | | | |
| Kole | 2022 | 31,9 | 23,4 | 16,5 | 13,5 | | | | | | |
| Lamwo | 2017 | 7,9 | 11,1 | 13,2 | 15,0 | 100,0 | 100,0 | 91,3 | 93,6 | 90,4 | 10,7 |
| Lamwo | 2018 | 37,9 | 31,7 | 27,0 | 22,9 | 100,0 | 100,0 | 96,6 | 93,6 | 85,5 | 10,0 |
| Lamwo | 2019 | 18,0 | 3,8 | 2,8 | 2,0 | 100,0 | 100,0 | 100,0 | 93,1 | 84,0 | 7,0 |
| Lamwo | 2020 | 1,4 | 1,2 | 67,7 | 68,6 | | | | 94,2 | 87,7 | 7,0 |
| Lamwo | 2021 | 70,3 | 70,6 | 71,2 | 34,2 | | | | | | |
| Lamwo | 2022 | 23,1 | 16,9 | 12,4 | 10,4 | | | | | | |
| Mubende | 2017 | 2,7 | 3,8 | 4,8 | 6,3 | 100,0 | 100,0 | 100,0 | 95,7 | 95,0 | 17,0 |
| Mubende | 2018 | 27,2 | 21,0 | 16,6 | 13,2 | 100,0 | 100,0 | 100,0 | 96,3 | 92,6 | 15,0 |
| Mubende | 2019 | 9,3 | 3,1 | 2,2 | 1,6 | 100,0 | 100,0 | 100,0 | 95,4 | 92,1 | 12,0 |
| Mubende | 2020 | 1,0 | 0,8 | 57,6 | 59,1 | | | | 95,8 | 93,9 | 9,8 |
| Mubende | 2021 | 61,9 | 63,2 | 64,5 | 52,0 | | | | | | |
| Mubende | 2022 | 32,4 | 22,7 | 15,5 | 12,0 | | | | | | |
| Rukiga | 2017 | 5,1 | 6,4 | 7,8 | 9,6 | 100,0 | 100,0 | 88,9 | 93,6 | 95,0 | 34,7 |

| District | Year | *pfcrt* K76T (%) | *pfmdr1* N86Y (%) | *pfmdr1* Y184F (%) | *pfmdr1* D1246Y (%) | *pfdhfr* S108N (%) | *pfdhfr* N51I (%) | *pfdhfr* C59R (%) | *pfdhps* A437G (%) | *pfdhps* K540E (%) | *pfdhps* A581G (%) |
|---|---|---|---|---|---|---|---|---|---|---|---|
| Rukiga | 2018 | 19,0 | 14,7 | 11,6 | 9,4 | | | | 93,5 | 92,7 | 29,5 |
| Rukiga | 2019 | 6,8 | 3,2 | 2,4 | 1,8 | 100,0 | 100,0 | 91,7 | 94,5 | 90,1 | 25,0 |
| Rukiga | 2020 | 1,2 | 1,0 | 51,3 | 52,8 | | | | 93,9 | 93,2 | 20,9 |
| Rukiga | 2021 | 55,5 | 56,8 | 58,1 | 26,6 | | | | | | |
| Rukiga | 2022 | 17,2 | 12,6 | 9,4 | 7,6 | | | | | | |
| Tororo | 2017 | 2,4 | 3,9 | 5,3 | 6,9 | 100,0 | 100,0 | 95,6 | 97,2 | 95,5 | 5,0 |
| Tororo | 2018 | 10,7 | 7,4 | 5,3 | 4,0 | 100,0 | 100,0 | 100,0 | 96,9 | 93,8 | 3,9 |
| Tororo | 2019 | 2,8 | 4,9 | 3,3 | 2,1 | 100,0 | 100,0 | 96,3 | 96,8 | 93,6 | 3,5 |
| Tororo | 2020 | 1,2 | 1,0 | 59,7 | 61,7 | 100,0 | 100,0 | 100,0 | 97,4 | 94,6 | 3,7 |
| Tororo | 2021 | 64,9 | 65,7 | 66,6 | 36,5 | 100,0 | 100,0 | 96,5 | | | |
| Tororo | 2022 | 23,9 | 17,4 | 12,6 | 10,2 | 99,1 | 99,2 | 95,1 | | | |

[1]Values are the district-year molecular-marker prevalence covariates contained in the final analytical dataset and used in bivariate regression screening. Variables stored as proportions in the analytical dataset (*pfcrt*, *pfmdr1* and *pfdhps*) are presented as percentages for readability. Blank cells indicate that the corresponding candidate molecular-marker covariate was unavailable for that district-year. Variables retained in the final multivariable model had complete data.

**Table S2. Bivariate associations between additional molecular resistance markers and malaria incidence in 16 Ugandan districts, 2017–2022[1]**

| Characteristic | Comparison or scale | Crude IRR | 95% CI | *p*-value |
|---|---|---|---|---|
| ***pfdhfr* markers** | | | | |
| *pfdhfr* S108N prevalence | Per 1-percentage-point increase | 0.501 | 0.307–0.816 | 0.005 |
| *pfdhfr* N51I prevalence | Per 1-percentage-point increase | 0.467 | 0.273–0.800 | 0.005 |
| *pfdhfr* C59R prevalence | Per 1-percentage-point increase | 1.004 | 0.995–1.012 | 0.400 |
| ***pfmdr1* markers** | | | | |
| *pfmdr1* N86Y prevalence[1] | Per 1.0 increase in proportion | 0.706 | 0.426–1.169 | 0.200 |
| *pfmdr1* Y184F prevalence | Per 1.0 increase in proportion | 0.925 | 0.559–1.531 | 0.800 |
| *pfmdr1* D1246Y prevalence | Per 1.0 increase in proportion | 1.038 | 0.617–1.745 | 0.900 |
| ***pfdhps* markers** | | | | |
| *pfdhps* A437G prevalence | Per 1.0 increase in proportion | <0.001 | <0.001–0.001 | 0.003 |
| *pfdhps* K540E prevalence | Per 1.0 increase in proportion | 0.000119 | 0.000001–0.026533 | 0.001 |
| *pfdhps* A581G prevalence | Per 1.0 increase in proportion | 27.606 | 1.926–395.735 | 0.015 |

[1]Molecular-marker prevalence (20) variables stored as proportions in the analytical dataset are reported on the scale used in the fitted models. The reported IRRs therefore correspond to a 1.0-unit increase in proportion, equivalent to a 100-percentage-point increase. The estimated IRR for *pfdhps* A437G was <0.000001 in the supplied regression output and is reported as <0.001 after rounding. IRR, incidence rate ratio; CI, confidence interval.

**Table S3. Model diagnostics and variable-selection assessment for the final negative binomial regression model[1]**

| Assessment | Variable / comparison | Result |
|---|---|---|
| **Model choice** | Dispersion parameter | 5.47 |
| | Regression family selected | Negative binomial |
| **Random effects** | Between-district SD | 0.648 |
| | Between-year SD | 0.652 |
| **Multicollinearity** | Combined *Kelch13* prevalence | VIF 1.21 |
| | ITN distribution | VIF 1.40 |
| | IRS coverage | VIF 1.15 |
| | Temperature | VIF 1.37 |
| | Relative humidity | VIF 1.36 |
| | Precipitation | VIF 1.09 |
| | Altitude | VIF 1.01 |
| | COVID-19 incidence | VIF 1.06 |
| | IPTp-SP3 uptake | VIF 1.14 |
| | *pfcrt* K76T | VIF 4.29 |
| | *pfmdr1* N86Y | VIF 4.66 |
| | *pfmdr1* D1246Y | VIF 1.56 |
| **Pairwise correlation** | *pfcrt* K76T vs *pfmdr1* N86Y | $r$=0.9801 |
| **Model comparison** | Final model retaining *pfcrt* K76T | AIC 8781.154 |
| | Alternative model retaining *pfmdr1* N86Y | AIC 8817.016 |
| **Selected marker** | Collinear predictor retained | *pfcrt* K76T |

[1]VIF, variance inflation factor; SD, standard deviation; AIC, Akaike Information Criterion; ITN, insecticide-treated net; IRS, indoor residual spraying; IPTp-SP3, intermittent preventive treatment in pregnancy with at least three doses of sulfadoxine-pyrimethamine. VIFs were assessed before multivariable modelling. Because *pfcrt* K76T and *pfmdr1* N86Y were strongly correlated, pairwise correlation and AIC were used alongside biological relevance to determine which predictor was retained in the final model. The model retaining *pfcrt* K76T had the lower AIC and was therefore selected.
